# Predicting the number of reported and unreported cases for the COVID-19 epidemic in South Korea, Italy, France and Germany

**DOI:** 10.1101/2020.03.21.20040154

**Authors:** P. Magal, G. Webb

## Abstract

We model the COVID-19 coronavirus epidemic in South Korea, Italy, France, and Germany. We use early reported case data to predict the cumulative number of reported cases to a final size. The key features of our model are the timing of implementation of major public policies restricting social movement, the identification and isolation of unreported cases, and the impact of asymptomatic infectious cases.

## 1 Introduction

In a previous work [2], we developed a model of the COVID-19 epidemic in China. The purpose of the model was to predict forward in time the future number of cases in a time-line of the epidemic from early reported case data. The model in [2] focused on the Chinese government imposed public policies designed to contain the epidemic. Our goal here is to apply this analysis to the COVID-19 epidemics in South Korea, Italy, France, and Germany. In an early phase of the epidemic, the reported case data grows exponentially, which corresponds to a constant transmission rate. We assume that government measures and public awareness cause this early constant transmission rate to change to a time dependent exponentially decreasing rate. We identify the early constant transmission rate using a method developed in [1]. We then identify the time dependent exponentially decreasing transmission rate from reported case data, and project forward the time-line of the epidemic course. We refer to [3, 4] for more results about this topic.

Our model incorporates the following essential ingredients of COVID-19 epidemics: (1) the number of asymptomatic infectious individuals (with very mild or no symptoms), (2) the number of symptomatic reported infectious individuals (with severe symptoms) and (3) the number of symptomatic unreported infectious individuals (with mild symptoms).

COVID-19 epidemics can be decomposed into three phases:

Phase I: *The first phase of the epidemic corresponds to a linear growth in the number of reported cases, where the number of daily reported case is almost constant day after day;*

Phase II: *The second phase of the epidemic corresponds to an exponential increasing phase, where the number of cases grows exponentially, corresponding to a constant transmission rate*.

Phase III: *The third phase of the epidemic corresponds to a time dependent exponentially decreasing transmission rate, due to major public interventions and social distancing measures*.

Our analysis identifies the epidemics in South Korea, Italy, France, and Germany as Phase III. Our model is applicable to COVID-19 epidemics in any region with reported case data, which can be updated to higher accuracy with on-going day by day reported case data.

## 2 Data

We use data from the Korean Center for Disease Control, the Italian Ministry of Health, the French Public Agency of Health, and the Robert Koch Institute of Germany. The cumulative reported cases are given in Figure 1 and the daily reported cases are given in Figure 2.

**Figure 1:**
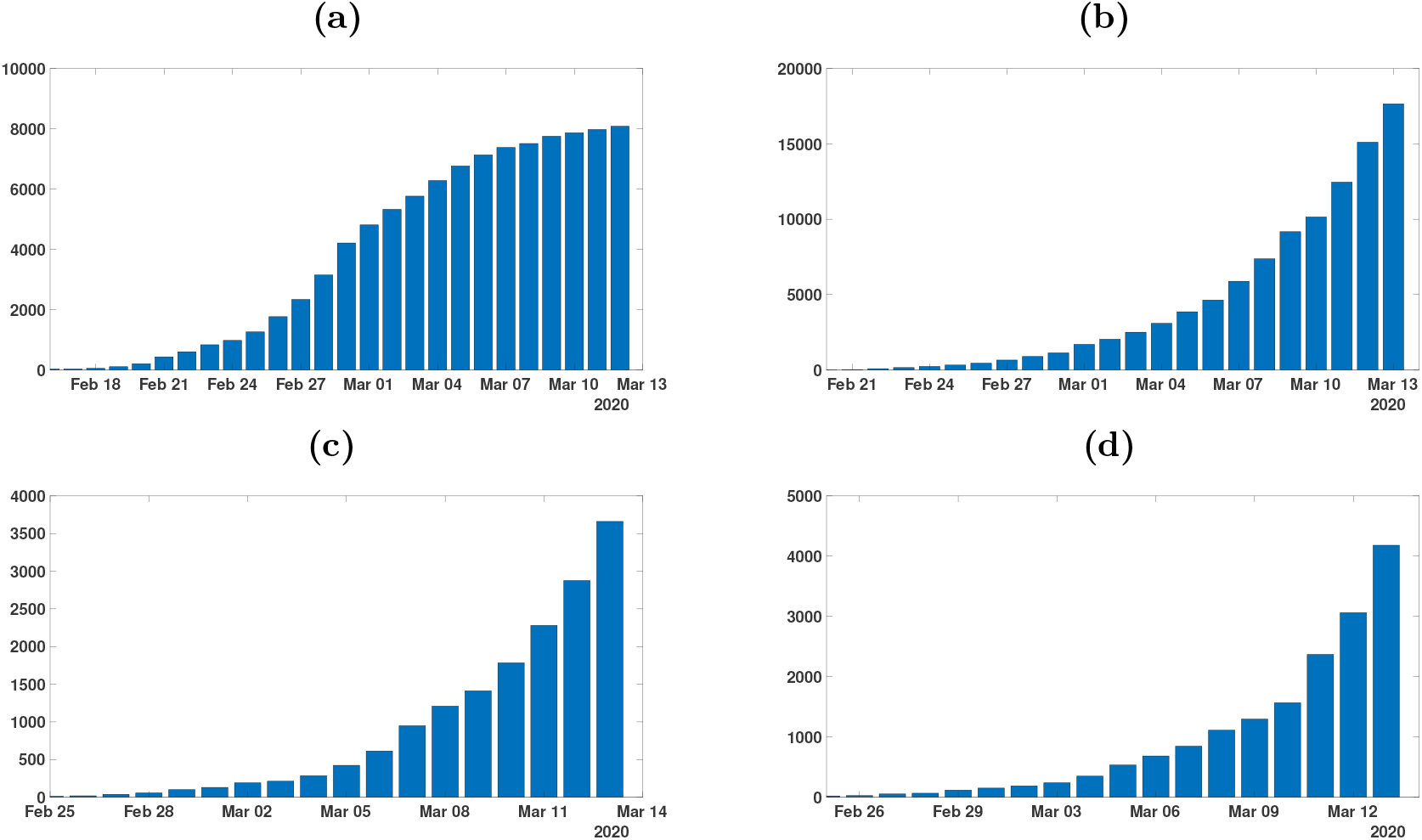
Cumulative number of reported cases for COVID-19 for (a) South Korea between January 20 and March 9; (b) Italy between January 31 and March 8; (c) France between February 25 and March 9; (d) Germany between February 24 and March 9.

**Figure 2:**
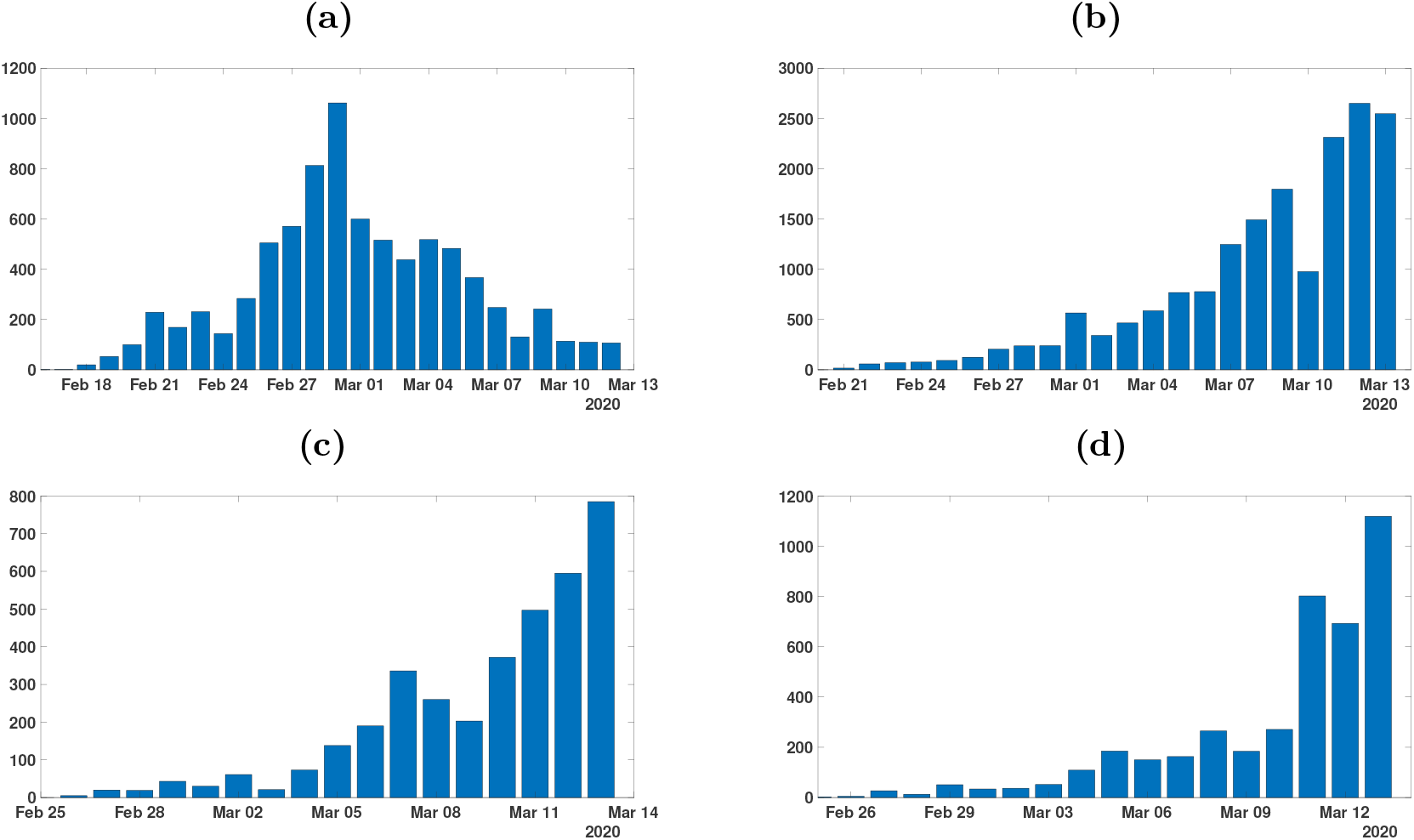
Daily number of reported cases for COVID-19 for (a) South Korea between January 20 and March 9; (b) Italy between January 31 and March 8; (c) France between February 25 and March 9; (d) Germany between February 24 and March 9.

## 3 Model

The model consists of the following system of ordinary differential equations:

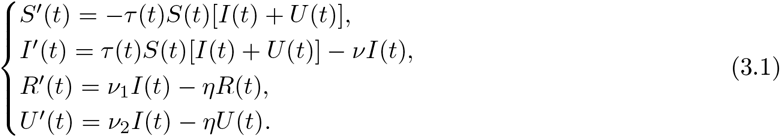

This system is supplemented by initial data

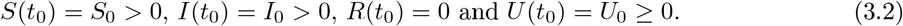

Here *t ≥ t*_0_ is time in days, *t*_0_ is the beginning date of the model of the epidemic, *S*(*t*) is the number of individuals susceptible to infection at time *t, I*(*t*) is the number of asymptomatic infectious individuals at time *t, R*(*t*) is the number of reported symptomatic infectious individuals at time *t*, and *U* (*t*) is the number of unreported symptomatic infectious individuals at time *t*.

The transmission rate at time *t* is *τ* (*t*). Asymptomatic infectious individuals *I*(*t*) are infectious for an average period of 1*/ν* days. Reported symptomatic individuals *R*(*t*) are infectious for an average period of 1*/η* days, as are unreported symptomatic individuals *U* (*t*). We assume that reported symptomatic infectious individuals *R*(*t*) are reported and isolated immediately, and cause no further infections. The asymptomatic individuals *I*(*t*) can also be viewed as having a low-level symptomatic state. All infections are acquired from either *I*(*t*) or *U* (*t*) individuals. The fraction *f* of asymptomatic infectious become reported symptomatic infectious, and the fraction 1 *− f* become unreported symptomatic infectious. The rate asymptomatic infectious become reported symptomatic is *ν*_1_ = *f ν*, the rate asymptomatic infectious become unreported symptomatic is *ν*_2_ = (1 *−f*) *ν*, where *ν*_1_ + *ν*_2_ = *ν*. The cumulative number of reported cases at time *t* is given by the formula

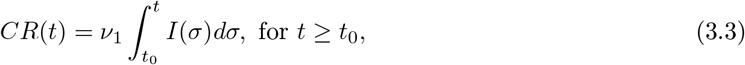

and the cumulative number of unreported at time *t* is given by the formula

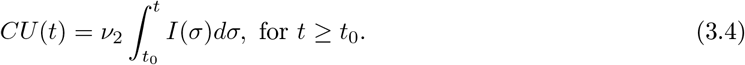

The parameters and initial conditions of the model are given in Table 1 and a flow diagram of the model is given in Figure 3.

**Table 1:** Parameters and initial conditions of the model.

| Symbol | Interpretation | Method |
| --- | --- | --- |
| $t_0$ | Time at which the epidemic started | fitted |
| $S_0$ | Number of susceptible at time $t_0$ | fixed |
| $I_0$ | Number of asymptomatic infectious at time $t_0$ | fitted |
| $U_0$ | Number of unreported symptomatic infectious at time $t_0$ | fitted |
| $\tau(t)$ | Transmission rate at time $t$ | fitted |
| $1/\nu$ | Average time during which asymptomatic infectious are asymptomatic | fixed |
| $f$ | Fraction of asymptomatic infectious that become reported symptomatic infectious | fixed |
| $\nu_1 = f\nu$ | Rate at which asymptomatic infectious become reported symptomatic | fitted |
| $\nu_2 = (1 - f)\nu$ | Rate at which asymptomatic infectious become unreported symptomatic | fitted |
| $1/\eta$ | Average time symptomatic infectious have symptoms | fixed |

**Figure 3:**
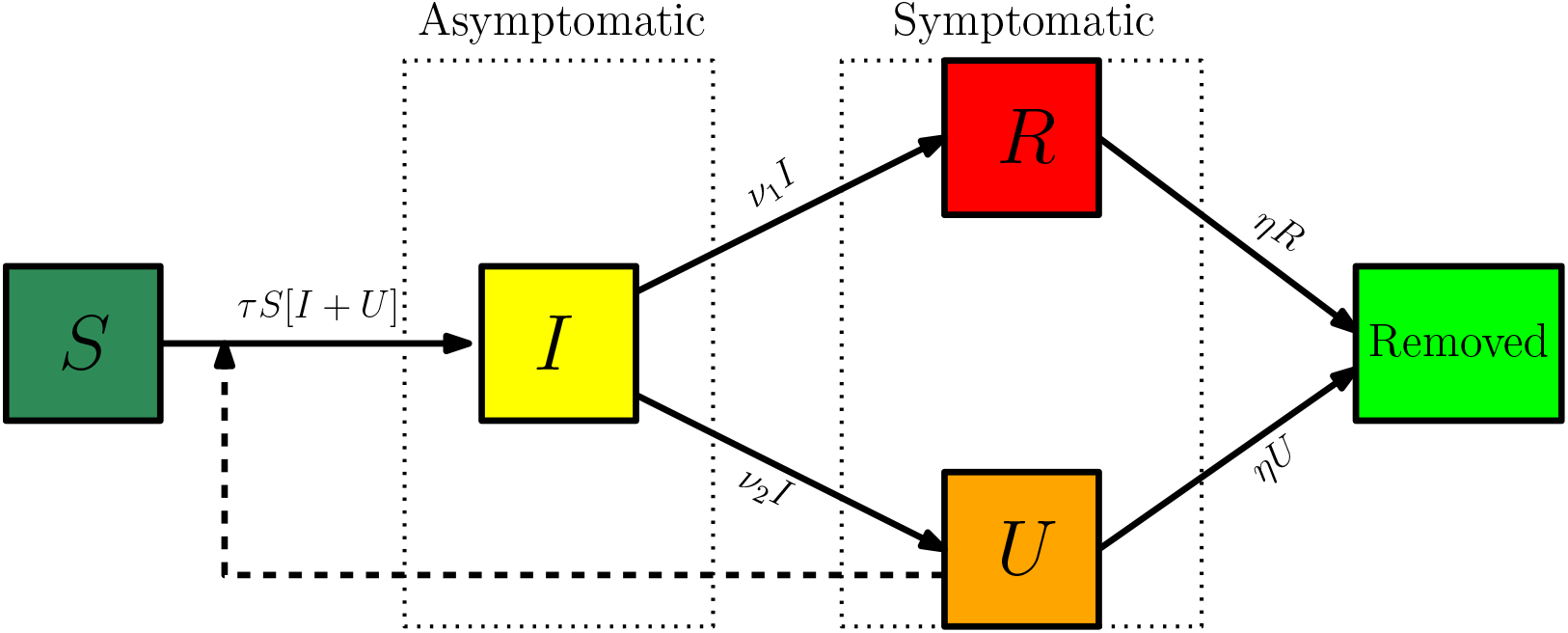
Compartments and flow chart of the model.

## 4 Method to estimate the parameters

We assume *f* = 0.6 or *f* = 0.1, which means that 40% or 90% of symptomatic infectious cases go unreported. The actual value of *f* is unknown. We assume *η* = 1*/*7, which means that the average period of infectiousness of both unreported symptomatic infectious individuals and reported symptomatic infectious individuals is 7 days. We assume *ν* = 1*/*7, which means that the average period of infectiousness of asymptomatic infectious individuals is 7 days. These values can be modified as further epidemiological information becomes known.

We assume that in the second phase of a COVID-19 epidemic, the cumulative number of reported cases *CR*(*t*) grows approximately exponentially, according to the formula:

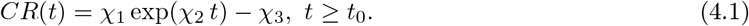

We fix the value *χ*_3_ = 1. The values of *χ*_1_ and *χ*_2_ are fitted to cumulative reported case data in the early phase of the epidemic, when it is recognized that *CR*(*t*) is growing exponentially (i.e. we use an exponential fit *χ*_1_ exp(*χ*_2_ *t*) to fit the data *CR*(*t*) + 1). We assume the initial value *S*_0_, corresponds to the population of the region of the reported case data. The value of the susceptible population *S*(*t*) is assumed to be only slightly changed by removal of the number of people infected in the beginning of the second phase. The other initial conditions are

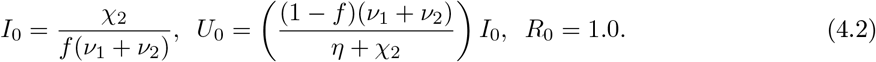

The value of the transmission rate *τ* (*t*), during the second phase of the epidemic, when the cumulative number of reported cases grows approximately exponential, is the constant value

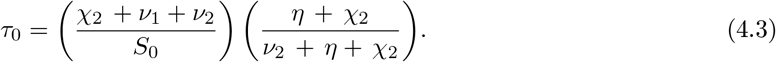

The initial time for the beginning of the early exponential growth phase is

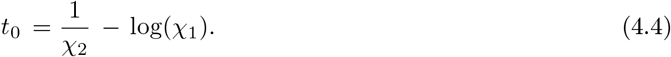

The value of the basic reproductive number is

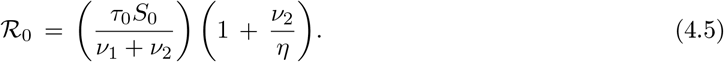

These formulas for *I*_0_, *U*_0_, *t*_0_, *τ*_0_, and *R*_0_ were derived in [1]. Their numerical values, which are technically theoretic, are essential for identification of the exponential growth rate of *CR*(*t*) in the second phase.

During the second phase *τ* (*t*)*≡ τ*_0_ is constant. When strong government measures such as isolation, quarantine, and public closings are implemented, the third phase begins. The actual effects of these measures are complex, and we use an exponential decrease for a time-dependent decreasing transmission rate *τ* (*t*) in the third phase to incorporate these effects. The formula for *τ* (*t*) during the third phase is

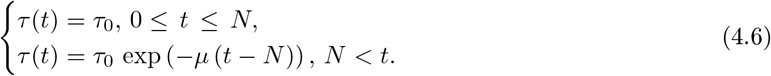

The date *N* and the value of *µ* are chosen so that the cumulative reported cases in the numerical simulation of the epidemic aligns with the cumulative reported case data after day *N*, when the public measures take effect. In this way we are able to project forward the time-path of the epidemic after the government imposed public restrictions take effect. We illustrate *τ* (*t*) in Figure 3 for a typical case.

The daily number of reported cases from the model can be obtained by computing the solution of the following equation:

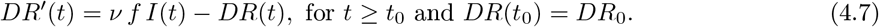

## 5 Predicting the cumulative number of cases

In this section we apply the method described in section 4 to the data coming from South Korea, Italy, France and Germany [5, 6, 7, 8]. In Table 2 we choose the data (1) February 19 to Feb 30 for South Korea; (2) February 21 to March 3 for Italy; (3) February 25 to March 7 for France; (4) February 24 to March 6 for Germany.

**Table 2:** The parameters χ_1_, χ_2_, χ_3_ are estimated by using the data in Figures 1 and 2 to fit χ_1_ exp(χ_2_ t) − χ_3_ to the data CR(t) between the following periods for each country: (1) February 19 to Feb 30 for South Korea; (2) February 21 to March 3 for Italy; (3) February 25 to March 7 for France; (4) February 24 to March 6 for Germany. The parameters ν = 1/7 and η = 1/7. The values of I_0_ U_0_, τ_0_, and t_0_ are obtained by using (4.2) to (4.4).

| Country | $\chi_1$ | $\chi_2$ | $\chi_3$ | $t_0$ | $\mu$ | $N$ | $S_0$ | $f$ | $\tau_0$ |
| --- | --- | --- | --- | --- | --- | --- | --- | --- | --- |
| South Korea | 0.624 | 0.294 | 1 | Feb. 2 | 1.3 | Feb 28 | $51.47 \times 10^6$ | 0.6 | $7.51 \times 10^{-9}$ |
| | | | | | | | | 0.1 | $6.56 \times 10^{-9}$ |
| Italy | 0.3459 | 0.2792 | 1 | Feb. 4 | 0.032 | Feb. 24 | $60.48 \times 10^6$ | 0.6 | $6.15 \times 10^{-9}$ |
| | | | | | | | | 0.1 | $6.56 \times 10^{-9}$ |
| France | 0.0020 | 0.3627 | 1 | Feb 17 | 0.032 | Feb. 28 | $66.99 \times 10^6$ | 0.6 | $6.78 \times 10^{-9}$ |
| | | | | | | | | 0.1 | $6.02 \times 10^{-9}$ |
| Germany | 0.0039 | 0.3371 | 1 | Feb 16 | 0.016 | Feb 28 | $82.79 \times 10^6$ | 0.6 | $5.18 \times 10^{-9}$ |
| | | | | | | | | 0.1 | $4.57 \times 10^{-9}$ |

### 5.1 Predicting the number of cases for South Korea

### 5.2 Predicting the number of cases for Italy

### 5.3 Predicting the number of cases for France

### 5.4 Predicting the number of cases for Germany

## 6 Conclusion

We have applied a method developed in [1] and [2] to predict the evolution of a COVID-19 epidemic in a geographical region, based of reported case data in that region. Our method uses early data, when the epidemic is in its second phase and growing exponentially, corresponding to a constant transmission rate. In [1] we demonstrated a method to identify this constant transmission rate. When public measures are begun in order to ameliorate the epidemic, a third phase begins, which we model with a time-dependent exponentially decreasing transmission rate in [2]. In [2] we applied this method to mainland China, and demonstrated the ability of our model to predict the forward time-line of the epidemic. In Figure 4 in [2], we showed how the prediction unfolded week by week, with increasing agreement with reported case data, in mainland China.

**Figure 4:**
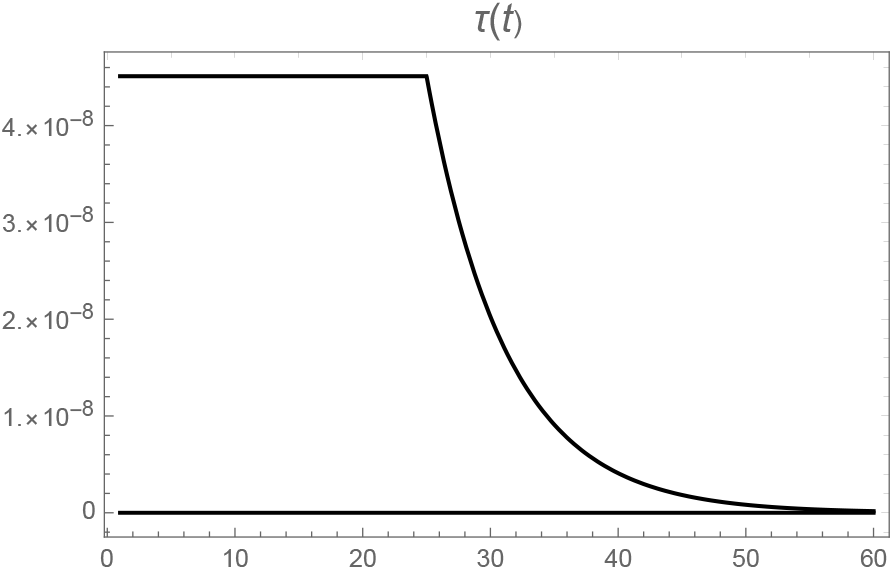
Graph of τ (t) with τ_0_ = 4.5× 10^−8^, N = 25, and µ = 0.16. The transmission rate is effectively 0 after day 53.

In this work we use the cumulative number of reported cases, the daily number of reported cases, and the weekly number of reported cases for four countries. With this data, we project the future number of cases, both reported and unreported, in each country. We assume a fraction *f* of total cases are unreported. We simulate our model with *f* = 0.6 and *f* = 0.1 for each country. The fractions *f* = 0.6 and *f* = 0.1 correspond to lesser and greater final sizes, respectively. For South Korea, the epidemic has subsided, because of the major measures that were implemented to restrict public distancing. For Italy, France, and Germany, the epidemics are still rising, because these measures have been implemented only recently.

Our model incorporates social distancing measures through the time dependent transmission rate *τ* (*t*) in the third phase. This rate involves *µ* (the rate of exponential decay corresponding to these measures) and *N* the day at which they become effective. It is evident that these measures should start as early as possible, and should be as strong as possible. The consequences of late public interventions may have severe consequences for the epidemic outcome (as illustrated in Figure 6 in [2]).

In the case of South Korea, the peak of the epidemic occurred approximately February 29. In Figures 5 and 6, we see that our model agrees very well the data for South Korea. Accordingly to our model, the daily number of cases reaches a maximum of approximately 700 cases near the turning point February 29.

**Figure 5:**
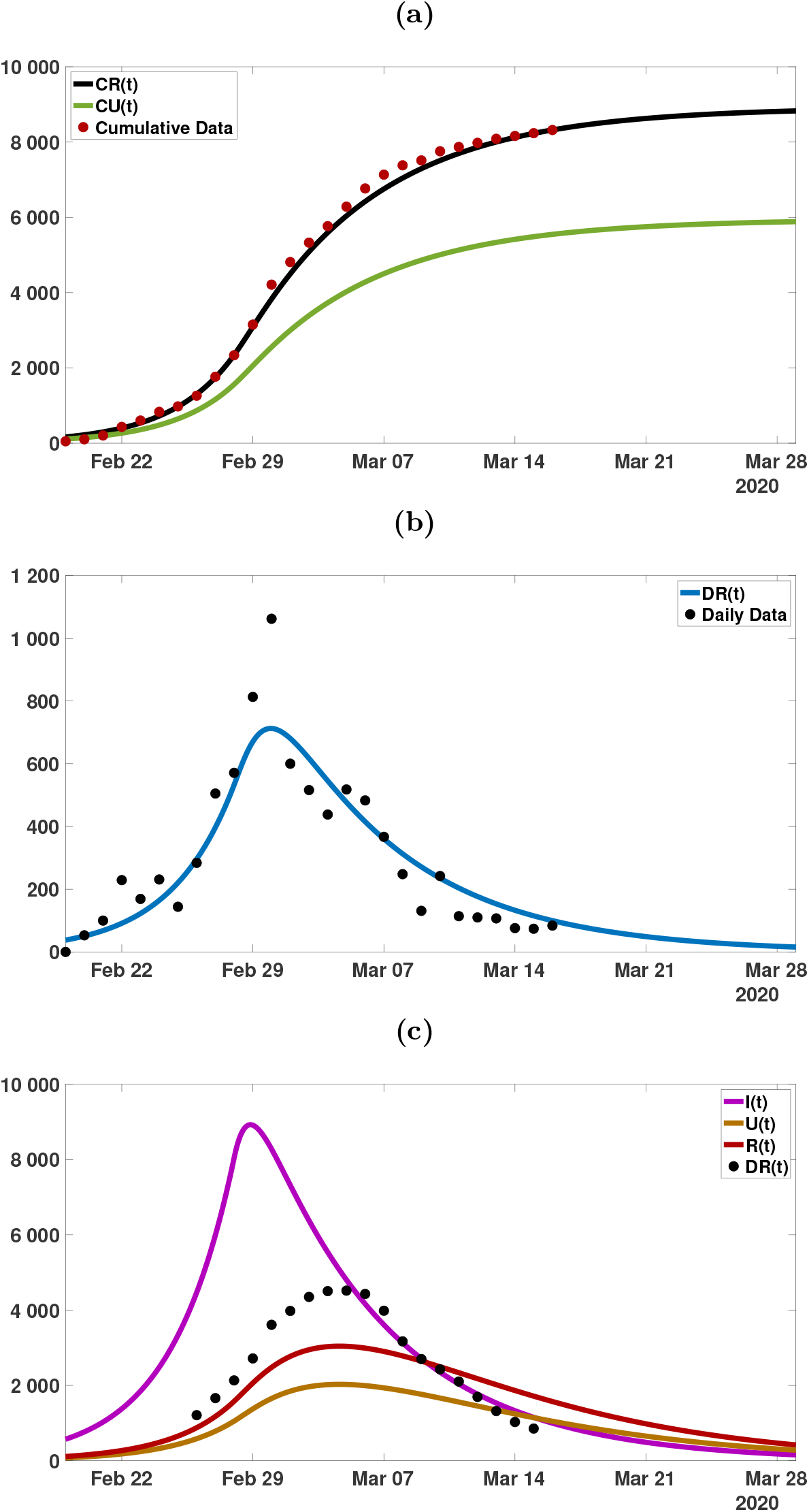
(a) Plot of the data for cumulative reported cases (red dots), CR(t) (black), and CU (t) (green) from the model, for f = 0.6 (60% of the cases are reported). I_0_ = 3.43, U_0_ = 0.45, ℛ_0_ = 3.79. The final size of the epidemic is approximately 8, 800 reported cases, and 5, 900 unreported cases. (b) Plot of the data (black dots) for the daily number of cases and DR(t) (blue) from the model. The turning point is approximately February 30. (c) Plot of I(t) (purple), U (t) (orange), R(t) (red) and the weekly data (blue dots) (each day minus 7 days earlier). The turning point of the weekly data and CR(t) and CU (t) is approximately March 4.

**Figure 6:**
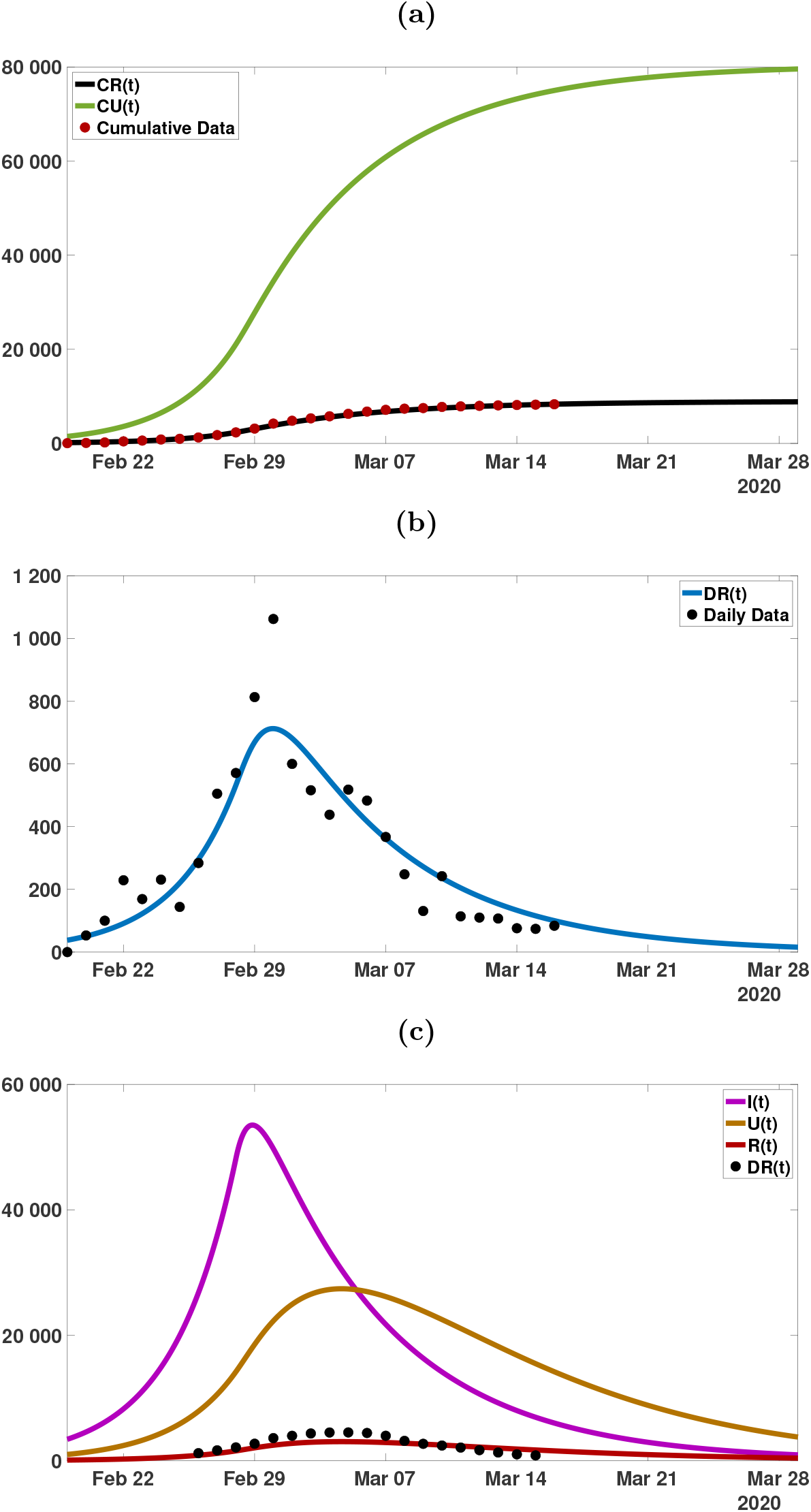
(a) Plot of the data for cumulative reported cases (red dots), CR(t) (black), and CU (t) (green) from the model, for f = 0.1 (10% of the cases are reported). I_0_ = 20.6, U_0_ = 6.1, ℛ_0_ = 4.49. The final size of the epidemic is approximately 10, 000 reported cases, and 80, 000 unreported cases. (b) Plot of the data (black dots) for the daily number of cases and DR(t) (blue) from the model. The turning point is approximately February 30. (c) Plot of I(t) (purple), U (t) (orange), R(t) (red) and the weekly data (blue dots) (each day minus 7 days earlier). The turning point of the weekly data and CR(t) and CU (t) is approximately March 4.

**Figure 7:**
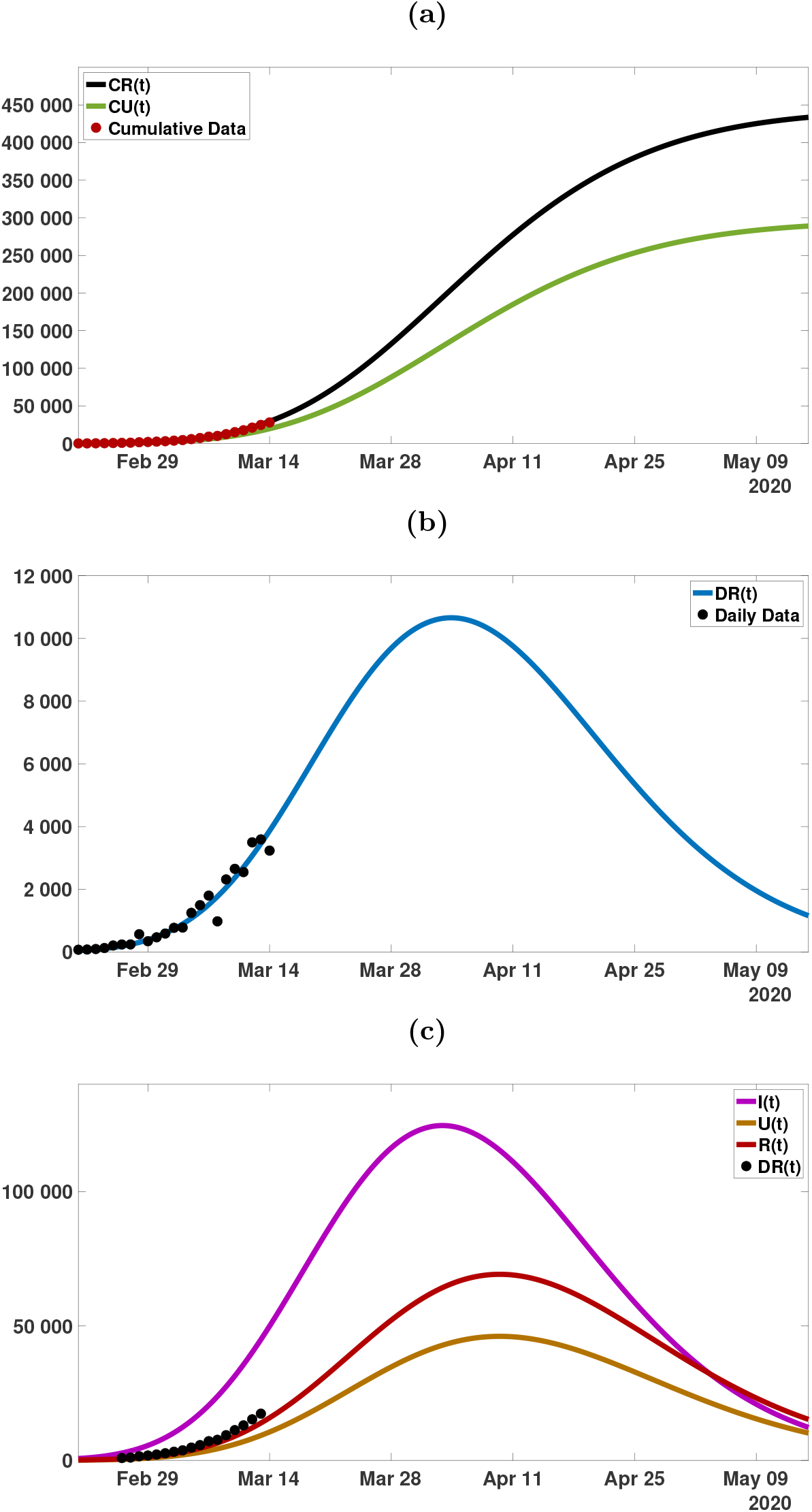
(a) Plot of the data for cumulative reported cases (red dots), CR(t) (black), and CU (t) (green) from the model, for f = 0.6 (60% of the cases are reported). I_0_ = 3.4, U_0_ = 0.45, ℛ_0_ = 3.79. The final size of the epidemic is approximately 435, 000 reported cases, and 280, 000 unreported cases. (b) Plot of the data (black dots) for the daily number of cases and DR(t) (blue) from the model. The turning point is approximately April 2. (c) Plot of I(t) (purple), U (t) (orange), R(t) (red) and the weekly data (blue dots) (each day minus 7 days earlier). The turning point of the weekly data and CR(t) and CU (t) is approximately March 31.

**Figure 8:**
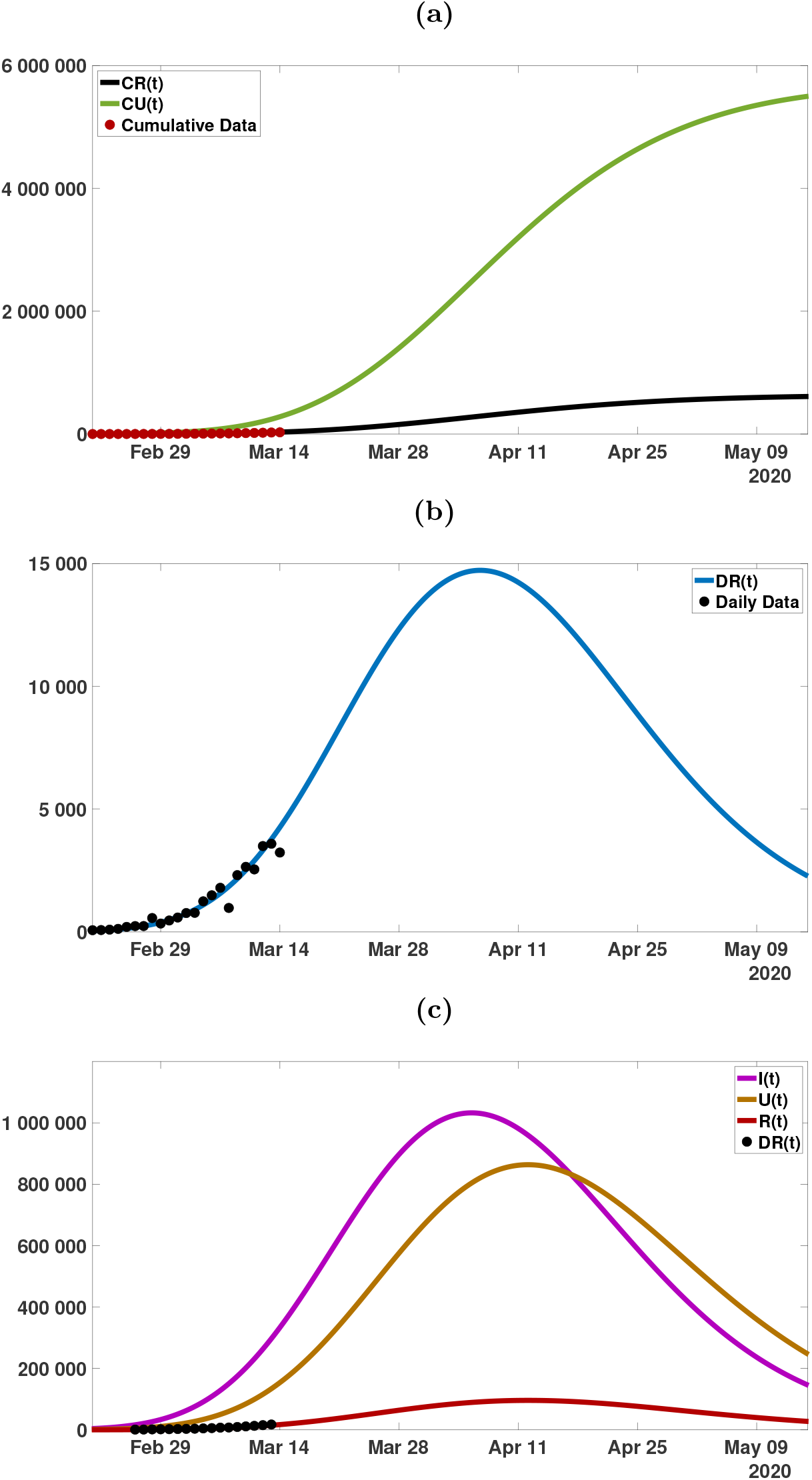
(a) Plot of the data for cumulative reported cases (red dots), CR(t) (black), and CU (t) (green) from the model, for f = 0.1 (10% of the cases are reported). I_0_ = 19.5, U_0_ = 6.0, ℛ_0_ = 4.30. The final size of the epidemic is approximately 600, 000 reported cases, and 5, 500, 000 unreported cases. (b) Plot of the data (black dots) for the daily number of cases and DR(t) (blue) from the model. The turning point is approximately April 7. (c) Plot of I(t)(purple), U (t) (orange), R(t) (red) and the weekly data (blue dots) (each day minus 7 days earlier). The turning point of the weekly data and CR(t) and CU (t) is approximately April 11.

**Figure 9:**
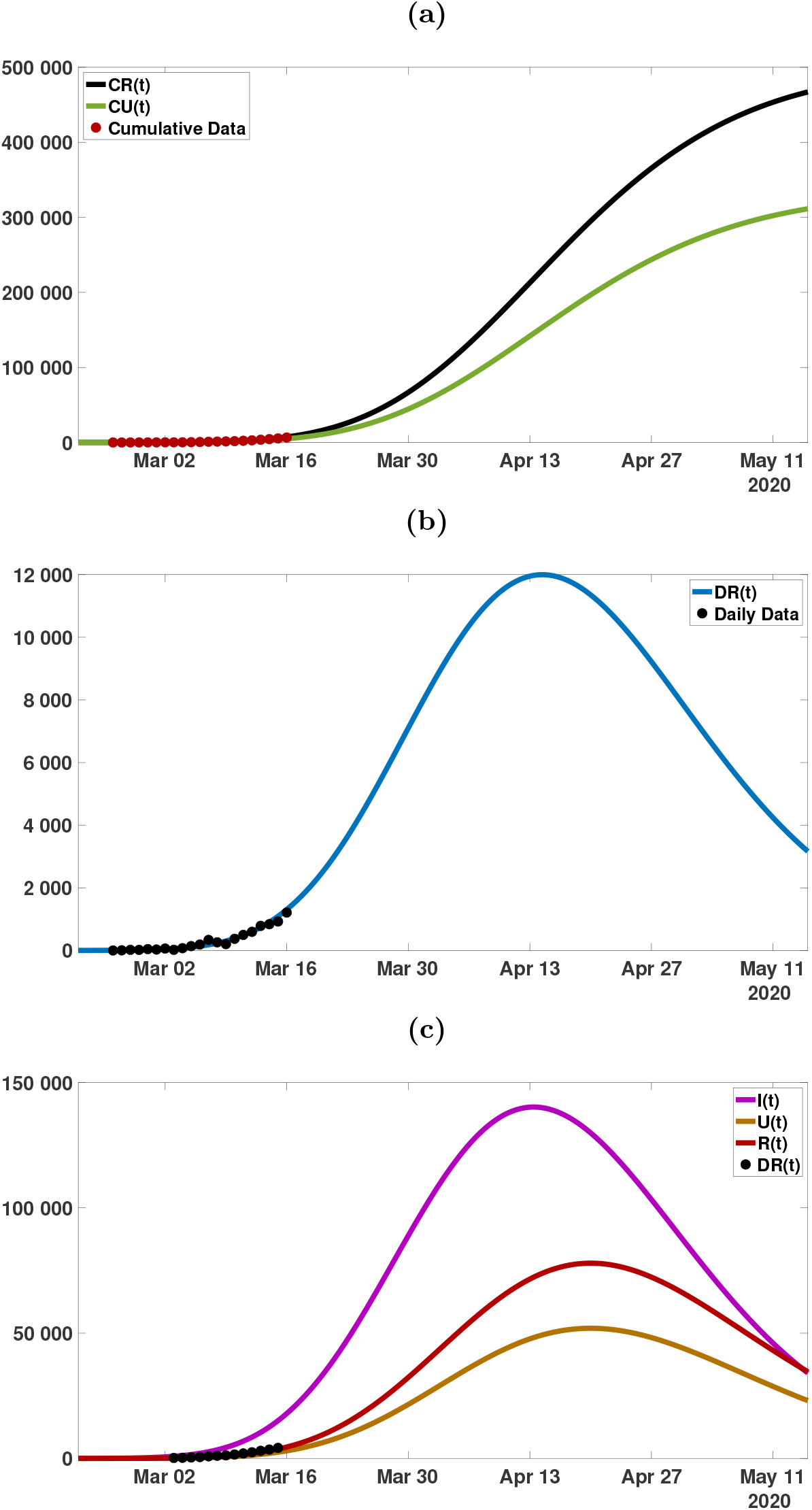
(a) Plot of the data for cumulative reported cases (red dots), CR(t) (black), and CU (t) (green) from the model, for f = 0.6 (60% of the cases are reported). I_0_ = 4.2, U_0_ = 0.58, ℛ_0_ = 4.45. The final size of the epidemic is approximately 470, 000 reported cases, and 310, 000 unreported cases. (b) Plot of the data (black dots) for the daily number of cases and DR(t) (blue) from the model. The turning point is approximately April 16. (c) Plot of I(t)(purple), U (t) (orange), R(t) (red) and the weekly data (blue dots) (each day minus 7 days earlier). The turning point of the weekly data and CR(t) and CU (t) is approximately April 20.

**Figure 10:**
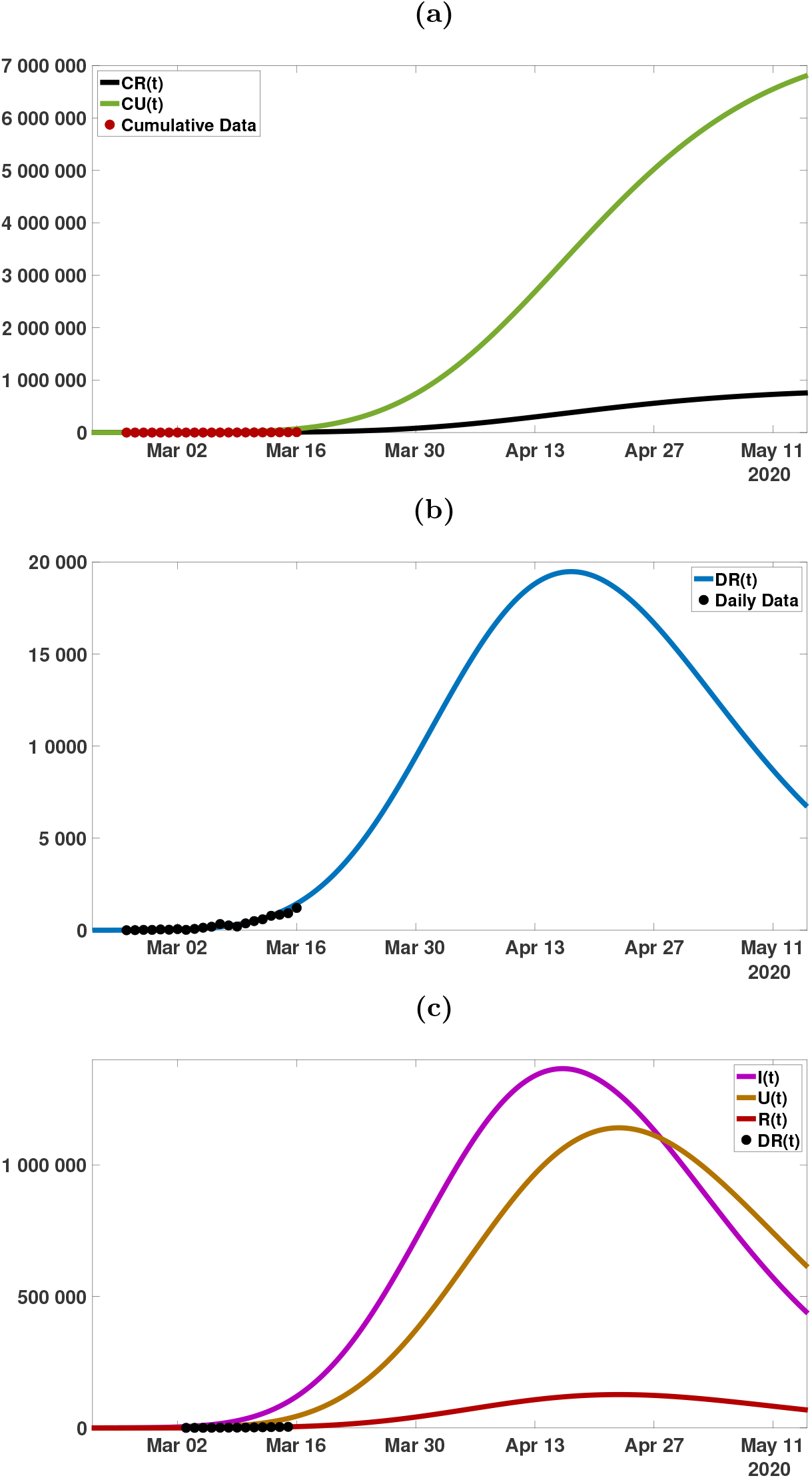
(a) Plot of the data for cumulative reported cases (red dots), CR(t) (black), and CU (t) (green) from the model, for f = 0.1 (10% of the cases are reported). I_0_ = 20.6, U_0_ = 6.1, ℛ_0_ = 4.49. The final size of the epidemic is approximately 700, 000 reported cases, and 6, 800, 000 unreported cases. (b) Plot of the data (black dots) for the daily number of cases and DR(t) (blue) from the model. The turning point is approximately April 17. (c) Plot of I(t)(purple), U (t) (orange), R(t) (red) and the weekly data (blue dots) (each day minus 7 days earlier). The turning point of the weekly data and CR(t) and CU (t) is approximately April 20.

**Figure 11:**
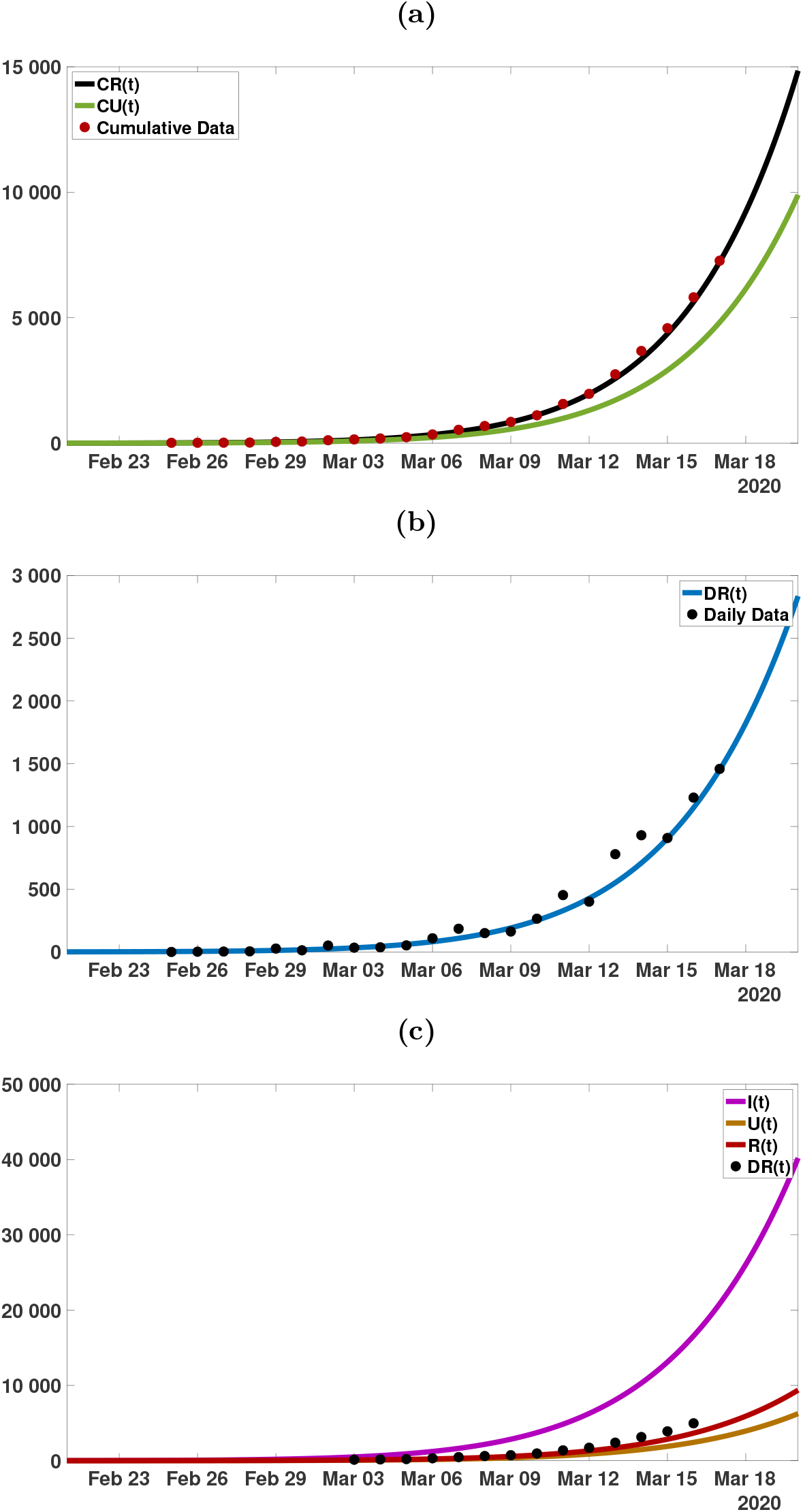
(a) Plot of the data for cumulative reported cases (red dots), CR(t) (black), and CU (t) (green) from the model, for f = 0.6 (60% of the cases are reported). I_0_ = 3.9, U_0_ = 0.47, ℛ_0_ = 4.21. (b) Plot of the data (black dots) for the daily number of cases and DR(t) (blue) from the model. (c) Plot of I(t)(purple), U (t) (orange), R(t) (red) and the weekly data (blue dots) (each day minus 7 days earlier).

**Figure 12:**
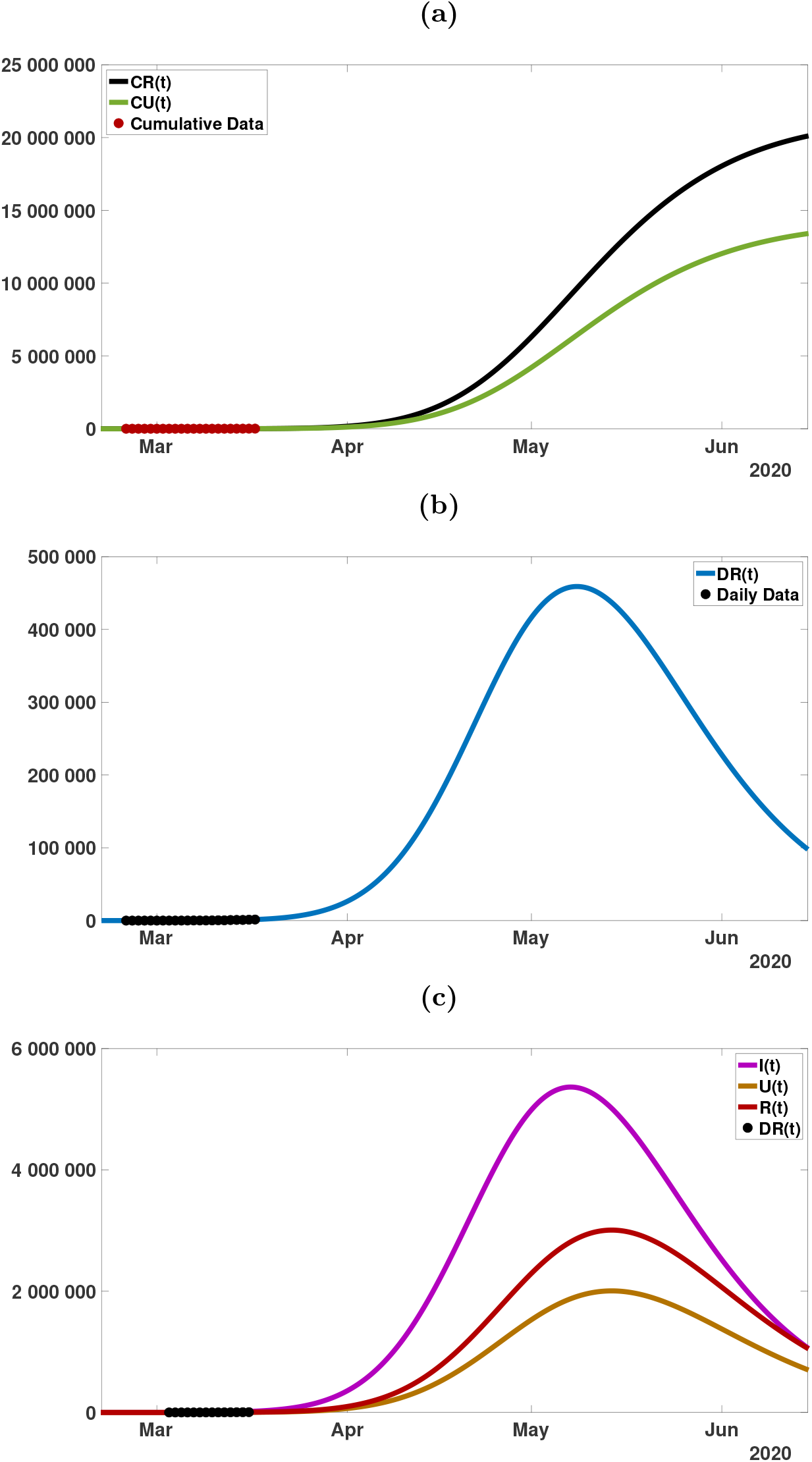
Graphs of the model simulation and data as in Figure 11, for a longer time period.

**Figure 13:**
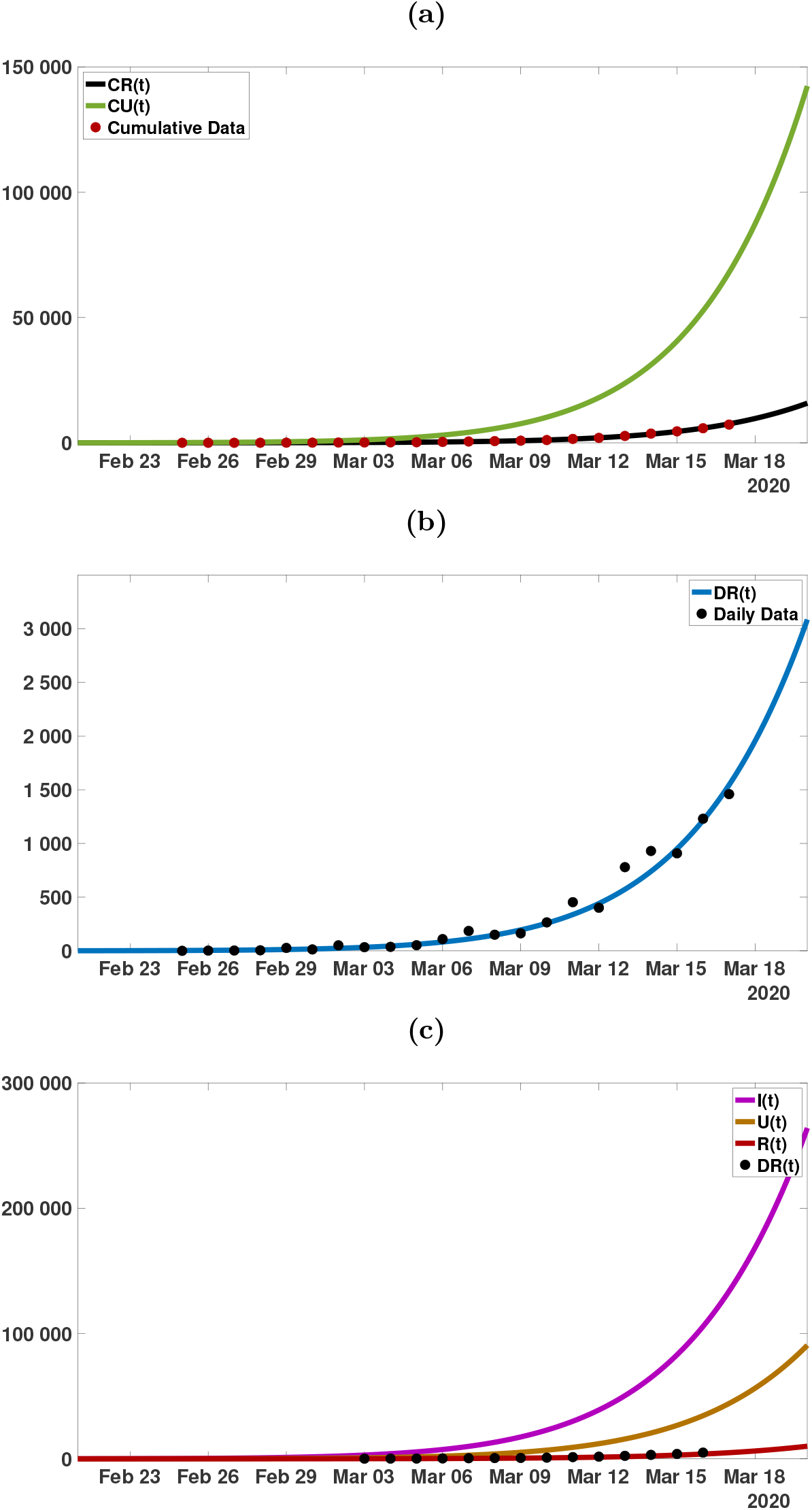
(a) Plot of the data for cumulative reported cases (red dots), CR(t) (black), and CU (t) (green) from the model, for f = 0.1 (10% of the cases are reported). I_0_ = 23.6, U_0_ = 6.3, ℛ_0_ = 5.03. (b) Plot of the data (black dots) for the daily number of cases and DR(t) (blue) from the model. (c) Plot of I(t)(purple), U (t) (orange), R(t) (red) and the weekly data (blue dots) (each day minus 7 days earlier).

**Figure 14:**
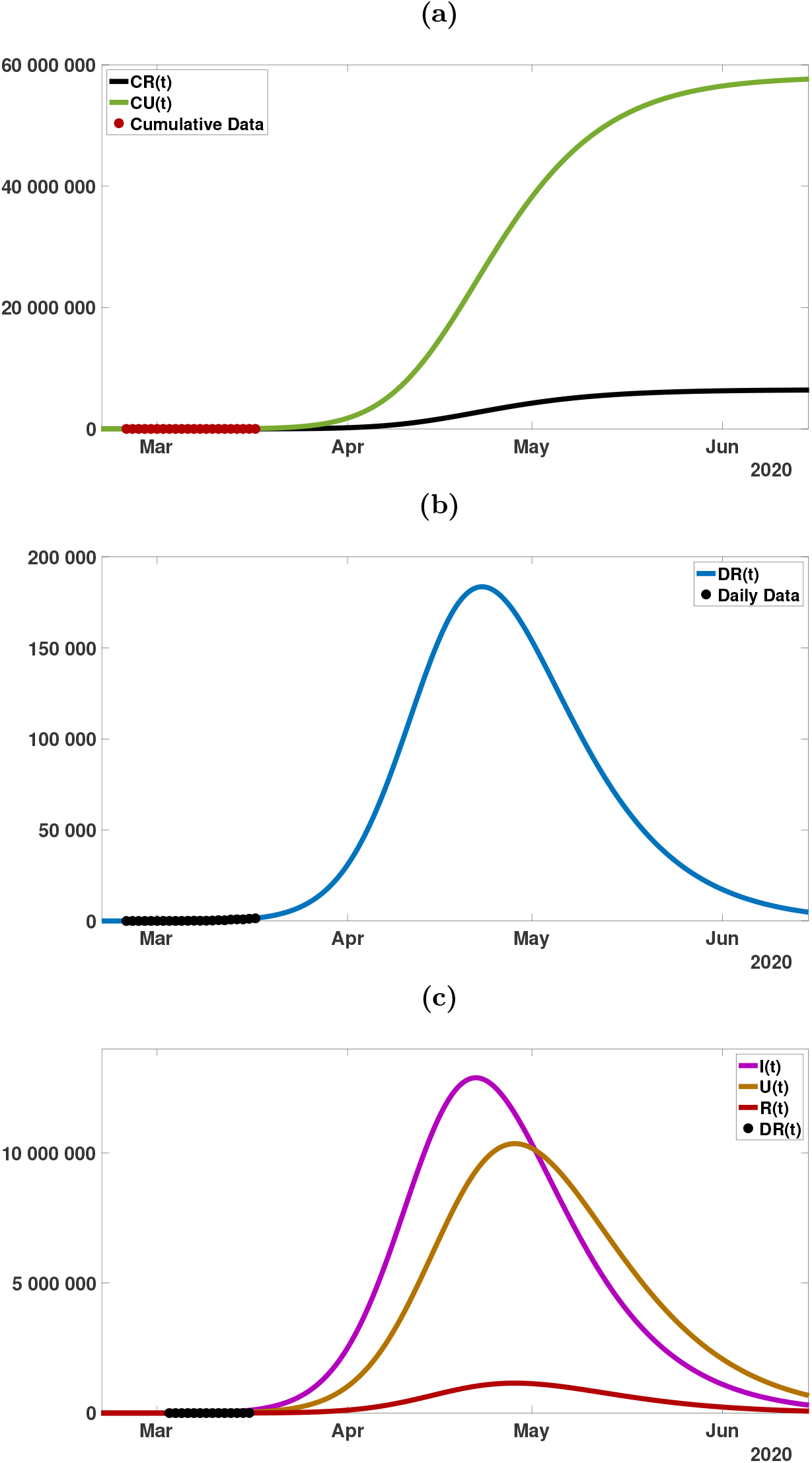
Graphs of the model simulation and data as in Figure 13, for a longer time period.

Compared to South Korea, the public interventions in Italy, France, and Germany were relatively late. The peak of the epidemic occurs in Italy around April 9, in France around April 14, and in Germany around May 1. The maximum daily number of cases is between 10 000 and 15 000 in Italy, between 12 000 and 20 000 in France, and between 200 000 and 500 000 in Germany. These numbers will likely remain high for an extended period of several months for these three countries, without further major public measures.

## Data Availability

The Data are coming from WHO.

## Conflicts of Interest

Declare conflicts of interest or state “The authors declare no conflict of interest.”

## Notes

### Competing Interest Statement

The authors have declared no competing interest.

### Funding Statement

No funding

